# Analysis of 16S rRNA gene sequence of nasopharyngeal exudate from healthy donors reveals changes in key microbial communities associated with aging

**DOI:** 10.1101/2022.06.26.22276913

**Authors:** Sergio Candel, Fernando Pérez-Sanz, Sylwia D. Tyrkalska, Antonio Moreno-Docón, Ángel Esteban, María L. Cayuela, Victoriano Mulero

**Affiliations:** Grupo de Inmunidad, Inflamación y Cáncer, Departamento de Biología Celular e Histología, Facultad de Biología, Universidad de Murcia, 30100 Murcia, Spain; Instituto Murciano de Investigación Biosanitaria (IMIB)-Arrixaca, 30120 Murcia, Spain; Centro de Investigación Biomédica en Red de Enfermedades Raras (CIBERER), Instituto de Salud Carlos III, 28029 Madrid, Spain; Servicio de Microbiología, Hospital Clínico Universitario Virgen de la Arrixaca, 30120 Murcia, Spain; Grupo de Telomerasa, Cáncer y Envejecimiento, Servicio de Cirugía, Hospital Clínico Universitario Virgen de la Arrixaca, 30120 Murcia, Spain

**Keywords:** Nasopharyngeal microbiome, Age differences, Sex differences, Aging, Human microbiome, 16s rRNA sequencing

## Abstract

**Background:** Functional or compositional perturbations of the microbiome can occur at different sites of the body and this dysbiosis has been linked to various diseases. Changes in the nasopharyngeal microbiome are associated to patient’s susceptibility to multiple viral infections, including COVID-19, supporting the idea that the nasopharynx may be playing an important role in health and disease. Most studies on the nasopharyngeal microbiome have focused on a specific component in the lifespan, such as infanthood or the elderly, or have other limitations such as low sample sizes. Therefore, detailed studies analyzing the age- and sex-associated changes in the nasopharyngeal microbiome of healthy people across their whole life are essential to understand the relevance of the nasopharynx in the pathogenesis of multiple diseases, particularly viral infections such as COVID-19.

**Results:** 120 nasopharyngeal samples from healthy subjects of all ages and both sexes were analyzed by 16s rRNA sequencing. Nasopharyngeal bacterial alpha diversity did not vary in any case between age or sex groups. Proteobacteria, Firmicutes, Actinobacteria, and Bacteroidetes were the predominant phyla in all the age groups, with several sex-associated differences probably due to the different levels of sex hormones between both sexes. *Acinetobacter, Brevundimonas, Dolosigranulum, Finegoldia, Haemophilus, Leptotrichia, Moraxella, Peptoniphilus, Pseudomonas, Rothia*, and *Staphylococcus* were the only 11 bacterial genera that presented significant age-associated differences. Other bacterial genera such as *Anaerococcus, Burkholderia, Campylobacter, Delftia, Prevotella, Neisseria, Propionibacterium, Streptococcus, Ralstonia, Sphingomonas*, and *Corynebacterium* appeared in the population with a very high frequency, suggesting that their presence might be biologically relevant.

**Conclusions:** In contrast to other anatomical areas such as the gut, bacterial diversity in the nasopharynx of healthy subjects remains very stable and resistant to perturbations throughout the whole life and in both sexes. Age-associated changes in taxonomic composition were observed at phylum, family, and genus levels, as well as several sex-associated changes probably due to the different levels of sex hormones present in both sexes at certain ages. Our results provide a complete and valuable dataset that will be useful for future research aiming for studying the relationship between changes in the nasopharyngeal microbiome and susceptibility to or severity of multiple diseases, including COVID-19.

## INTRODUCTION

Among the remaining challenges in biomedical sciences, one of the most important is to fully understand how aging affects human biological processes, health, and wellness. In recent years, solid evidence has been found to support the idea that the microbial communities that inhabit the different anatomical areas of the human body could play a key role in these processes, and there has been much speculation about possible medical interventions ^1-6^. Although most research has focused on the well-studied gut microbiome, there is growing evidence that variations in microbial communities in other sites of the body are also responsible for wide-ranging health effects ^7-11^. The case of the respiratory tract is curious, since the lungs were long believed to be sterile, despite the fact that they are constantly exposed to microorganisms in inhaled air and the upper respiratory tract ^12^, which has been the main cause that the respiratory microbiome has barely been studied until very recently ^13^. However, new culture-independent microbial identification techniques, such as metagenomics, have revealed that the respiratory tract is a dynamic ecosystem and has raised the interest of the scientific community in the role of the respiratory microbiota in health and disease ^12,13^.

The human upper respiratory tract, that comprises the anterior nares, nasal cavity, sinuses, nasopharynx, Eustachian tube, middle ear cavity, oral cavity, oropharynx, and larynx, is the major portal of entry for infectious droplet or aerosol-transmitted microorganisms ^14^. Among these different areas, the nasopharynx is anatomically unique because it presents a common meeting place for the ear, nose, and mouth cavities ^15^, but has not gained special prominence until the outbreak of the current COVID-19 pandemic ^16^. Nasopharyngeal swabs from COVID-19 patients present higher viral loads and diagnostic sensitivities than oropharyngeal ^17^, anterior nares, ^18^ and throat ^19^ swabs, suggesting the relevance of the nasopharyngeal epithelium as a portal of initial infection and transmission of SARS-CoV-2. Thus, the nasopharynx has become the main anatomical area from which samples are collected for SARS-CoV-2 viral testing worldwide. Importantly, dozens of studies have already found unquestionable correlations between the composition of the nasopharyngeal microbiota and susceptibility to different viral infections in humans ^13^, and there is beginning to be some evidence, although still controversial, that it may be playing a role in the susceptibility to SARS-CoV-2 infection too ^20,21^. Elucidating this might shed light on the still unexplained fact that some COVID-19 patients, such as the elderly, are more susceptible and present more severe forms of COVID-19 than others ^22^.

Large cohort studies of human microbiome data with appropriate controls are particularly valuable, especially of all ages and both sexes, as these datasets are difficult to obtain due to multiple factors, including our long lifespans, heterogeneity in consent and other sample access issues, and because of socioeconomic confounds. Therefore, human studies have tended to focus on a specific component of the lifespan, such as infanthood, or studies of the elderly, rather than examining variation across an entire population. Knowledge about the relationships between changes in the nasopharyngeal microbiota and susceptibility to viral infections is a good example of this, since most studies have focused only on children ^13^. Interestingly, the case of COVID-19 is not an exception, and the controversy over whether changes in the nasopharyngeal microbiome affect disease severity at different ages is probably because most studies present important limitations, such as low sample sizes or focusing only on a specific life stage. Thus, while some studies revealing changes in the nasopharyngeal microbiome of COVID-19 patients do not present appropriate uninfected controls ^21^ or have been performed only in children ^20^, others that do not find any relevant microbiome alterations have low sample sizes ^23,24^.

A crucial factor to consider in studies of variation across the lifespan is sex as a biological variable. For aging research, this includes the understanding that females and males may have different aging trajectories ^25-27^, including in key systems such as the digestive tract. For example, the gut microbiome and sex hormones may interact to predispose women to autoimmune diseases ^28^ and dietary interventions are known to have sex-specific effects on gut microbiota ^29^. There are no studies analyzing the possible sex-associated differences in the nasopharyngeal microbiome at different life stages.

Here, we analyze in 120 healthy individuals of all ages and both sexes, for the first time, the diversity and taxonomic composition of the nasopharyngeal microbiota across the whole lifespan, the taxonomic changes in the nasopharynx associated to age or sex, and the possible biological relevance of several taxa whose frequency of appearance in the population is high. We therefore provide a very comprehensive and valuable dataset, that will be the base for future research aimed at identifying relationships between age- and sex-associated changes in nasopharyngeal microbiome and susceptibility to or severity of the diseases of interest, including COVID-19.

## RESULTS

### Data annotation and samples overview

A total of 120 nasopharyngeal microbiomes from 120 healthy individuals were analyzed. A total of 4,538,196 high-quality 16S rRNA sequences ranging from 10,627 to 256,449 sequences per sample (mean = 37,818.3; median = 33,169) were obtained after quality control analyses and OUT filtering. The 16S rRNA sequences were binned into 128 families, 250 genera and 561 species. The most abundant families were Staphylococcaceae (12.14%), Burkholderiaceae (11.52%), Carnobacteriaceae (11.48%) and Corynebacteriaceae (9.47%). The most abundant genera were *Staphylococcus* (13.06%), *Dolosigranulum* (11.99%), *Corynebacterium* (10.18%) and *Ralstonia* (10.08%). The most abundant species were *Dolosigranulum pigrum* (24.55%), *Ralstonia pickettii* (19.02%), *Corynebacterium pseudodiphtheriticum* (4.87%) and *Propionibacterium acnes* (4.85%). We excluded one sample with an abnormally high proportion of *Chlamydophila*, which is an indication of an abnormal sampling or pathological disorder of this individual ‘C_A1_M8’ (data not shown). To reveal age-related progression of nasopharyngeal microbiota, we divided the samples into 6 age groups, each divided into females and males to be able to also study possible sex-associated differences (Table S1). There were 20 samples in each age group and 10 samples in each sex group within them, except for the first age group (A1: 1-20 years) where one male had to be excluded as indicated above (Table S1).

We performed Principal Component Analysis (PCA) to visualize the taxonomic patterns of these samples in a low-dimension space based on the relative abundance matrix of the 250 genera across the 119 samples. Thus, the top two principal components explained 17.68% and 10% of the variance of the original data, respectively (Fig. 1). As shown in Fig. 1, the samples from individuals younger than 41 and older than 70 years were scattered loosely, while the rest of samples appeared mostly intermixed. However, the samples did not form distinct clusters when viewed using this linear approach. Something similar was found when focusing on possible differences between the two sexes, as samples from females and males also appeared completely intermixed and did not form any groups (Fig. 1).

**Figure 1.**
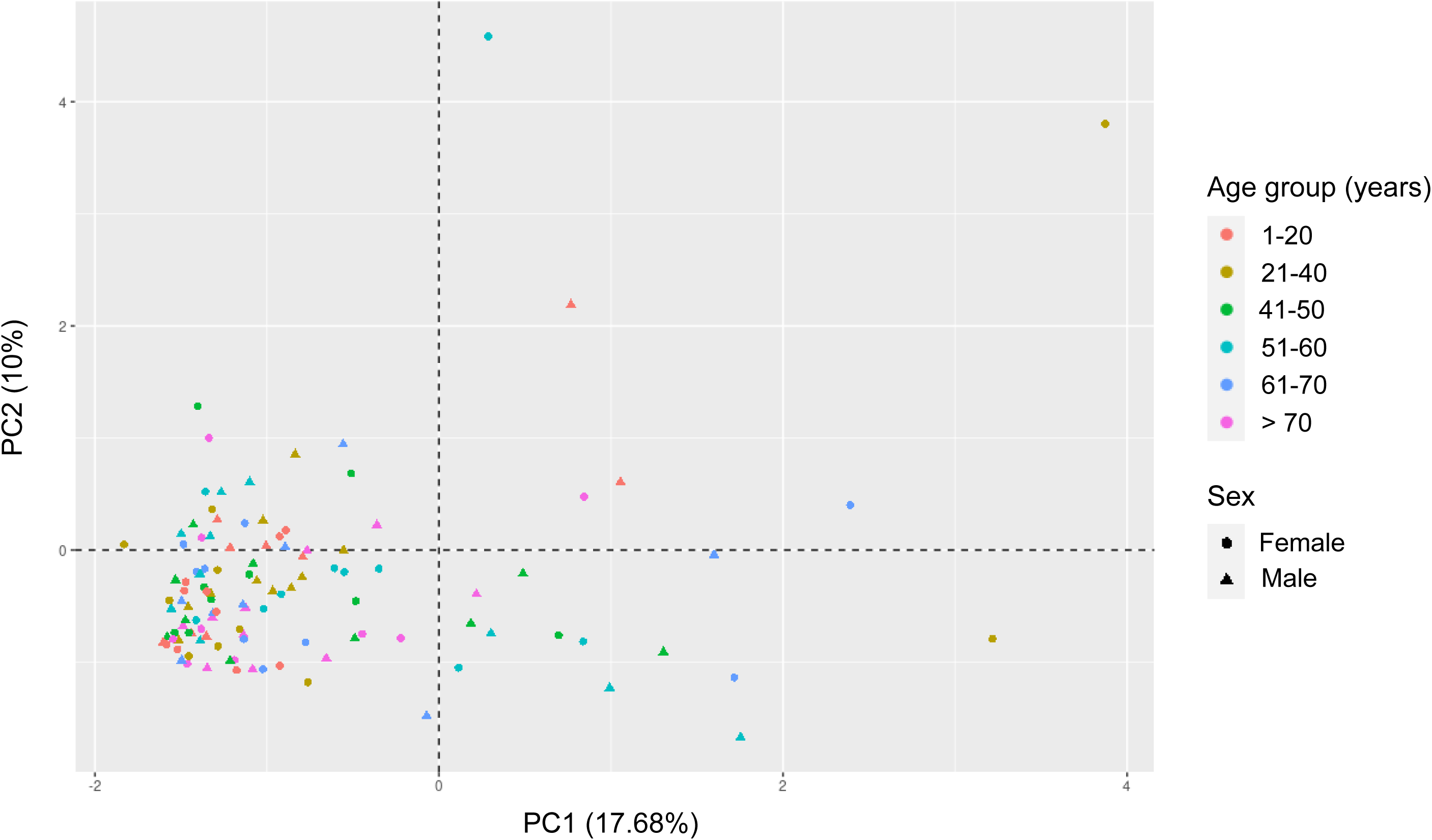
Sample overview using PCA. Using the relative abundance of 250 genera in the 119 samples as input, the data were linearly transformed and visualized in two-dimensional space. Each sample is represented by one dot, colored according to age, and shaped according to sex. The percentage of the variance of the original data explained by each of the two principal components is indicated in the axis labels.

### Bacterial diversity in the nasopharynx of healthy individuals is stable throughout lifespan

The fact that significant changes in bacterial diversity throughout life had previously been described in the well-studied gut microbiota of healthy individuals ^30^, prompted us to test whether similar changes occur in the nasopharynx by analyzing the alpha diversity, referred to as within-community diversity ^31^, for the different age and sex groups established for this study (Table S1). However, the Shannon’s diversity index, which measures evenness and richness of communities within a sample, did not show any statistically significant changes in bacterial diversity among the different age groups (Fig. 2a). Moreover, alpha diversity also did not vary as a function of sex when the Shannon index was calculated considering all individuals of all ages included in this study (Fig. 2b), nor when the same analysis was performed comparing females and males within each age group (Fig. 2c). The use of other indexes commonly used to measure alpha diversity, such as the inverse Simpson’s diversity index, which is an indication of the richness in a community with uniform evenness that would have the same level of diversity (Fig. S1), or the Chao1 index, which measures the total richness of communities within a sample (Fig. S2), confirmed the absence of any statistically significant differences in bacterial diversity between the different age groups (Fig. S1a and S2a) or between females and males (Fig. S1b-c and S2b-c). Therefore, all these results together suggest that contrary to what occurs in other anatomical areas, such as the gut where bacterial diversity decreases with aging ^30^, it remains stable in the nasopharynx of healthy people over time, without notable changes at any stage of life, not even in very young people or in the elderly over 70 years of age (Fig. 2a, S1a and S2a). Curiously, another interesting finding provided by this work, for the first time, is that there are no significant differences when comparing bacterial diversity in the nasopharynx of healthy females and males (Fig. 2b-c, S1b-c, and S2b-c), regardless of the stage of life studied and the important hormonal differences that exist between both sexes at certain ages.

**Figure 2.**
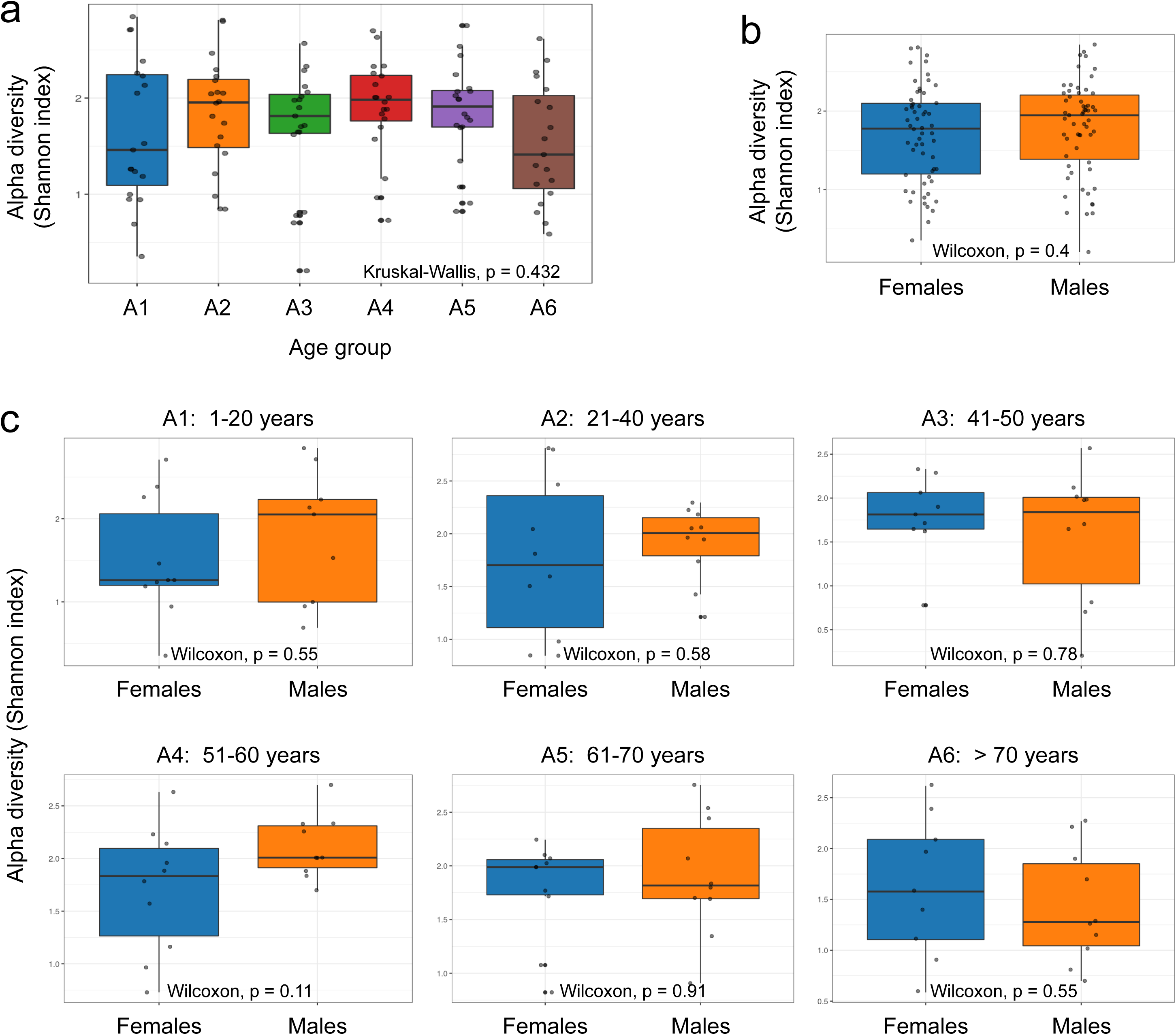
Comparison of alpha diversity parameters across the age and sex groups studied. Box-whisker plots of the alpha diversity Shannon index and its comparison using the Kruskal-Wallis test among the different age groups established for this study (**a**), and the Wilcoxon signed-rank test between females and males (**b-c**). Each sample is represented by one dot.

### Aging- and sex-associated changes in compositional structure of nasopharyngeal microbiome in healthy individuals

To determine differences in nasopharyngeal taxonomic composition among age and sex classes, we compared the nasopharyngeal microbiome of healthy women and men in the six different age groups established for this study (Table S1). A first general analysis at the phylum level revealed that Firmicutes and Proteobacteria relative abundances showed opposite kinetics with aging (Fig. 3a-f): while the relative abundance of Firmicutes, which is the majority phylum in the youngest individuals (50%) (Fig. 3a), clearly decreased with aging reaching its lowest values in subjects in their 50’s and 60’s (21% and 27%, respectively) (Fig. 3d-e), the relative abundance of Proteobacteria presented its lowest value in the youngest people (24%) (Fig. 3a) and increased in older individuals, peaking in subjects who are in their 50’s (53%) (Fig. 3d). The relative abundance of other phyla, such as Actinobacteria, Bacteroidetes, Tenericutes, or Cyanobateria, remained more stable throughout life (Fig. 3a-f). Moreover, when we continued working at the phylum level and looked for any differences between the nasopharyngeal taxonomic composition of females and males, we found that, interestingly, the results were almost identical for both sexes within the two age groups containing the youngest (1-20 years) (Fig. S3a) and oldest (> 70 years) (Fig. S3f) individuals. However, several differences between females and males were observed in other age groups, notably a higher relative abundance of Actinobacteria in males in their 20s and 30s compared to females of the same age group (20% vs. 7%) (Fig. S3b), a higher relative abundance of Proteobacteria in males in their 40s compared to females of the same age group (46% vs. 29%) (Fig. S3c), a higher relative abundance of Proteobacteria in females in their 50s compared to males of the same age group (57% vs. 47%) (Fig. S3d), and a higher relative abundance of Firmicutes and lower of Actinobacteria in females in their 60s compared to males of the same age group (37% vs. 20% and 10% vs. 22%, respectively) (Fig. S3e). Therefore, the fact that there were differences between the nasopharyngeal taxonomic composition of females and males in most of the age groups studied (individuals between 21 and 70 years of age) (Fig. S3b-e), but not within the two age groups containing the youngest and oldest individuals (people between 1 and 20 and older than 70 years of age) (Fig. S3a and f), suggests that the different levels of sex hormones present in both sexes at different life stages might be modulating the taxonomic composition of the nasopharyngeal microbiome.

**Figure 3.**
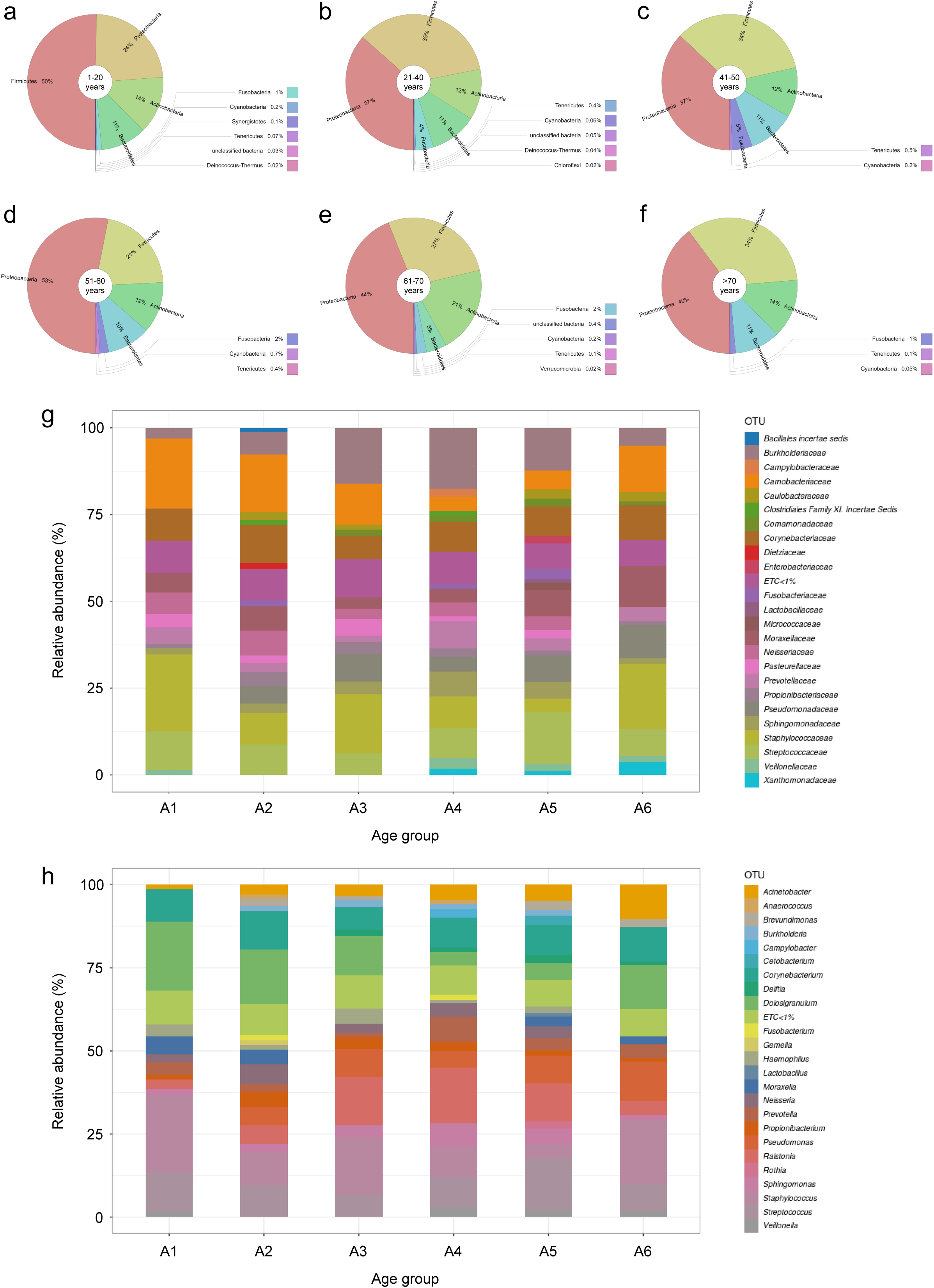
Taxonomic composition and age-associated metagenomic changes in the nasopharynx of healthy individuals. **a-f** Krona charts showing the bacterial community composition at the phylum level in the indicated age groups. Stacked bar charts showing the relative abundance (%) of bacterial phyla. **g** Stacked bar charts showing the relative abundance (%) of bacterial families in the indicated age groups. **h** Stacked bar charts showing the relative abundance (%) of bacterial genera in the indicated age groups. For clarity, only bacterial families (**g**) and genera (**h**) with average abundance > 1% at each age group are shown.

Next, we wanted to go further in this study and moved to the family level, finding that 24 distinct bacterial families presented an average abundance > 1% in at least one of the age groups studied (Fig. 3g). Our analyses revealed which was the dominant family in each of the age groups: Staphylococcaceae in A1, Carnobacteriaceae in A2, Staphylococcaceae in A3, Burkholderiaceae in A4, Streptococcaceae in A5, and Staphylococcaceae in A6 (Fig. 3g). Thus, it is curious that although Proteobacteria was the majority taxa in all the age groups when analyzing the nasopharyngeal taxonomic composition at the phylum level (Fig. 3a-f), the dominant family in all the age groups belongs to the Firmicutes phylum, except in the case of age group A4 where the family Burkholderiaceae, which belongs to the Proteobacteria phylum, was the most abundant taxa at family level (Fig. 3g). Differences in the relative abundance of some families were found when compared between age groups, but without following any easily interpretable pattern (Fig. 3g). Analysis of the nasopharyngeal taxonomic composition at the family level between age groups, but separately in females and males, did not show any relevant sex-associated differences between sexes regarding the dominant bacterial families in each age group compared to the results described above for both sexes together (Fig. 3g), excepting that Burkholderiaceae was the most abundant family in males of age group A3 instead of Staphylococcaceae, and that Corynebacteriaceae is the dominant family in males of age group A5 instead of Streptococcaceae (Fig. S4). As mentioned above when working with females and males together, differences in relative abundance were found in some families when comparing between different age groups in both sexes, but without following any easily interpretable patterns (Fig. S4). Visualization of the nasopharyngeal taxonomic composition at the family level of all the individuals included in this study showed that most of them had a very diverse microbiome, with a high number of families with relative abundance > 1% (Fig. S5). However, a few individuals had one dominant family that represented the majority of their nasopharyngeal microbiomes; these families tended to be Burkholderiaceae, Carnobacteriaceae and Staphylococcaceae (Fig. S5).

Working at the genus level, we found that 24 bacterial genera presented an average abundance > 1% in at least one of the age groups studied (Fig. 3h). Moreover, our results showed that *Staphylococcus* was the dominant genus in age group A1, *Dolosigranulum* in A2, *Staphylococcus* in A3 and A6, *Ralstonia* in A4, and *Streptococcus* in A5 (Fig. 3h). No relevant differences were found when comparing the nasopharyngeal taxonomic composition of females and males at the genus level within the age groups studied (Fig. S6). Similar to what was described above at the family level, visualization of the nasopharyngeal taxonomic composition at the genus level of all individuals included in this study showed that most of them had a high number of genera with relative abundance > 1%, with a few individuals presenting one dominant genus (mostly *Dolosigranulum* or *Staphylococcus*) that represented the majority of their nasopharyngeal microbiomes (Fig. S7). Next, we focused on those genera whose relative abundance were significantly different between the different age groups established in this study, as this could help us to identify changes in the nasopharyngeal taxonomic composition that are characteristic of aging. Thus, our analyses revealed that there were statistically significant differences (adjusted p-value < 0.05) in relative abundance between the distinct age groups in 11 bacterial genera: *Acinetobacter, Brevundimonas, Dolosigranulum, Finegoldia, Haemophilus, Leptotrichia, Moraxella, Peptoniphilus, Pseudomonas, Rothia* and *Staphylococcus* (Fig. 4a and Table S2). Interestingly, most of these statistically significant differences in relative abundance between the age groups for the 11 mentioned genera were between age groups A1 or A6, which include individuals between 1 and 20 years of age and over 70 years old, respectively, and the rest of age groups (18 out of 37 cases for the age group A1 and 16 out of 37 cases for the age group A6) (Table S2). Among these statistically significant changes found, it should be noted that *Acinetobacter* was the only genus whose relative abundance in the nasopharynx clearly increased progressively throughout life, peaking in individuals older than 70 years of age (Fig. 4a and Table S2). In the cases of *Dolosigranulum* and *Rothia*, their relative abundance drastically increased and decreased, respectively, in individuals over 70 years of age, compared to middle-aged people in their 50s and 60s (Fig. 4a and Table S2). Changes among the different age groups of *Finegoldia, Leptotrichia* and *Haemophilus* were also interesting, as their relative abundance was markedly reduced in elderly people over 70 years of age, even though they were present at other ages throughout life, mainly during middle age (Fig. 4a and Table S2). The case of *Haemophilus* was particularly intriguing, as while its relative abundance was at least 10% of the nasopharyngeal microbiota composition in age groups A1-A5 (if we consider only these 11 genera that present statistically significant differences between age groups), it dramatically decreased in the group of individuals over 70 years old (Fig. 4a and Table S2). Furthermore, it is noteworthy that the bacterial genera *Brevundimonas, Finegoldia, Leptotrichia* and *Peptoniphilus* presented a very low relative abundance in the youngest individuals, who are between 1 and 20 years old, compared to other age groups (Fig. 4a and Table S2). Finally, although *Moraxella* and *Staphylococcus* showed significant differences in relative abundance between the distinct age groups in several cases, these differences did not seem to follow any easily interpretable pattern relating relative abundance levels to a particular life stage (Fig. 4a and Table S2). Next, we wondered whether the significant differences in relative abundance between age groups observed for these 11 bacterial genera were due to sex-associated differences. Visualization of the nasopharyngeal taxonomic composition of females and males separately, considering only these 11 genera, showed no notable differences between both sexes (Fig. S8). The only exception was a higher relative abundance of *Dolosigranulum* in males in their 20s and 30s compared to females of the same age, because this genus was clearly dominant in 5 males from that age group while only in 1 female of the same age (Fig. S9). Besides this observation regarding *Dolosigranulum*, analyzing the taxonomic composition of all the individuals included in this study also revealed that in most people, 1 out of these 11 genera was dominant compared to the relative abundance of the other 10 genera (Fig. S9). Interestingly, 8 out of the 11 genera, excepting *Bevundimonas, Finegoldia* and *Peptoniphilus*, were found to be the dominant genus in at least 1 individual, demonstrating that nasopharyngeal taxonomic composition at this level can be very variable between different individuals, even if they belong to the same age or sex groups (Fig. S9 and S10).

**Figure 4.**
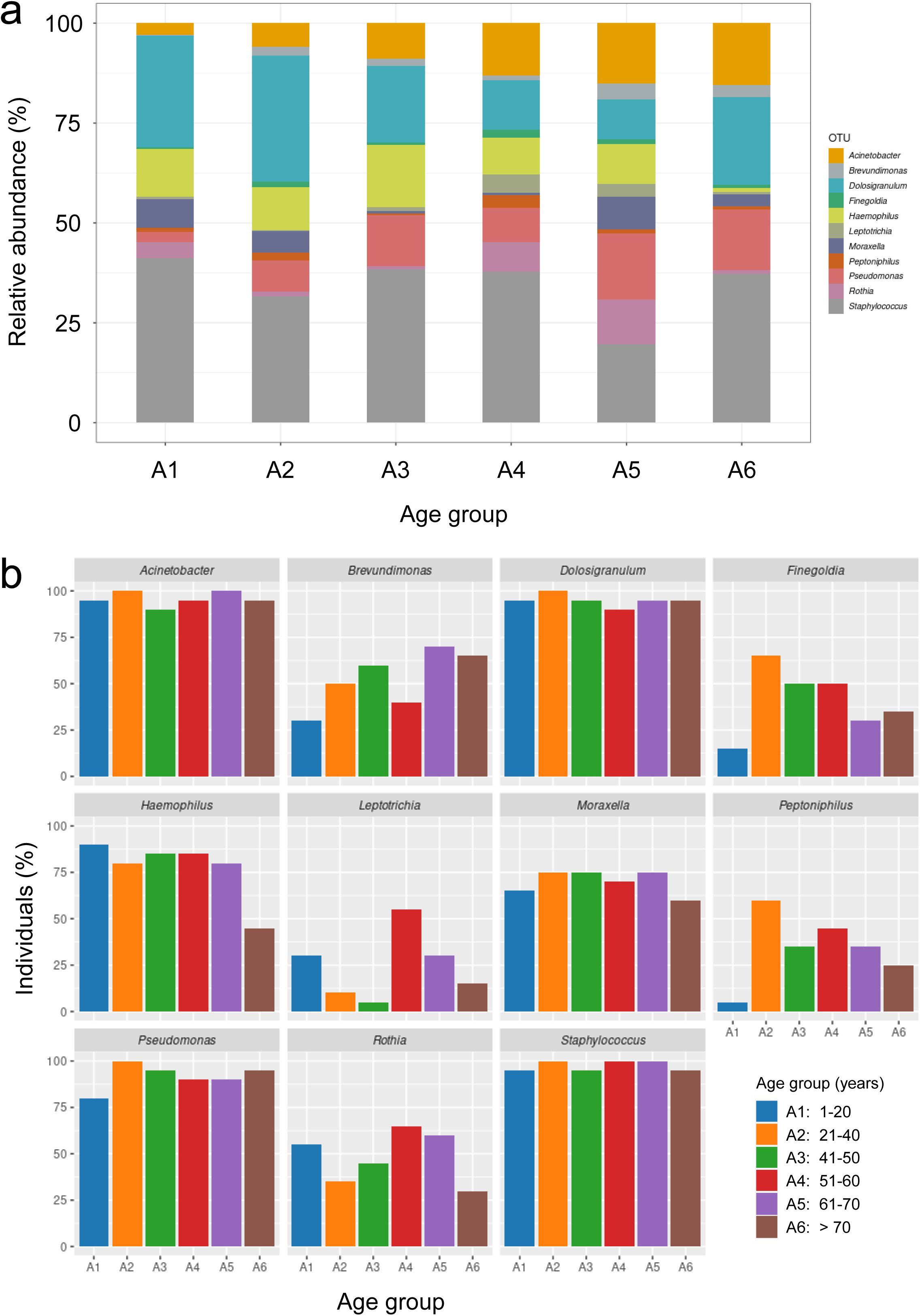
Taxonomic composition and frequency of appearance of the 11 bacterial genera which show significant differences between age groups. **a** Stacked bar charts showing the relative abundance (%) of the 11 bacterial genera indicated in the age groups established for this study. **b** Percentage of individuals, of the total included in this study, in which the indicated genera are present in the indicated age groups.

### Identification of potentially biologically relevant bacterial genera by analyzing their frequency of appearance in the nasopharynx of healthy individuals

After analyzing the nasopharyngeal taxonomic composition at the phylum, family, and genus levels, looking for differences in relative abundance between age and sex groups, we decided to apply another strategy to try to identify bacterial genera whose presence in the nasopharynx might be characteristic of a certain life stage, independently on their abundance levels. Thus, based on the idea that in some cases the presence of a genus within a certain sex or age group but not in others can be biologically relevant, even if its abundance is low, we studied the frequency with which each genus appeared in the individuals included in this study by visualizing, for each genus, the percentage of people from each age group in which it is present. Firstly, we analyzed this in the 11 genera that presented significant differences between the distinct age groups, with the aim of checking whether these genera were present in a high percentage of individuals and, therefore, that the relative abundance results previously shown in Fig. 4a were reliable. Indeed, our results showed that these 11 genera appear with a high frequency in the individuals included in this study, especially in the cases of *Acinetobacter, Dolosigranulum, Haemophilus, Moraxella, Pseudomonas* and *Staphylococcus* (Fig. 4b). These data also revealed that several genera, such as *Brevundimonas, Finegoldia* and *Peptoniphilus* appeared less frequently in the youngest people than in the other age groups, while the frequency of appearance of others, such as *Rothia*, decreases in the oldest people (Fig. 4b). Interestingly, it coincides that these genera that appear most frequently in the individuals included in this study are also the ones that appear most often as the dominant genus in the taxonomic composition of individuals (Fig. S9 and S10). Next, we identified several bacterial genera that could be biologically relevant as part of the nasopharyngeal microbiota although they did not show any significant differences in relative abundance between age or sex groups. This was (i) because their frequency of appearance in the nasopharynx of the healthy population was very high, as in the case of *Anaerococcus, Burkholderia, Campylobacter, Delftia, Prevotella, Neisseria, Propionibacterium, Streptococcus, Ralstonia, Sphingomonas* and *Corynebacterium* (Fig. 5a); (ii) because their frequency of appearance drastically increased with age, such as the cases of *Faecalibacterium, Stenotrophomonas* and *Phascolarctobacterium* (Fig. 5b); or (iii) because their frequency of appearance drastically decreased with age, such as the cases of *Aggregatibacter, Gemella* and *Fusobacterium* (Fig. 5c).

**Figure 5.**
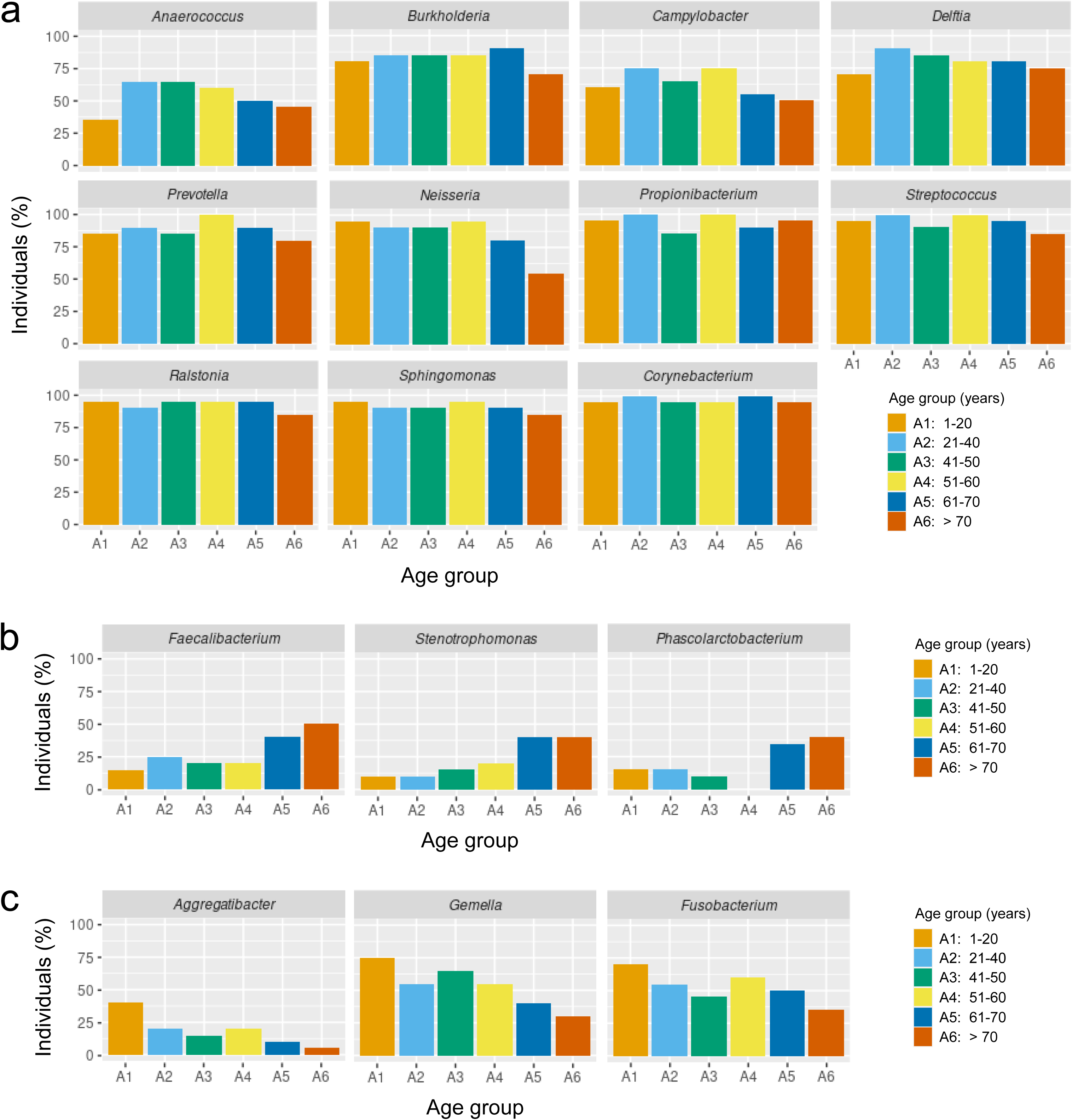
Frequency of appearance of potential biologically relevant bacterial genera. **a-c** Percentage of individuals, of the total included in this study, in which the indicated genera are present in the indicated age groups.

## DISCUSSION

Comparing bacterial species richness in the nasopharynx of healthy individuals in the different age groups established for this study and between females and males revealed no age- or sex-associated significant differences in alpha diversity. These results, which were confirmed by using three of the most reliable and commonly used alpha diversity indexes, such as the Shannon’s diversity index, the inverse Simpson’s diversity index, and the Chao1 diversity index, indicate that the nasopharyngeal microbiome is highly stable and robust to perturbations throughout life as well as between sexes within each of the different age groups in healthy subjects. Alpha diversity could be expected to be lower in the youngest individuals compared to older individuals in all the anatomical areas, as the fetus is sterile until the moment of birth, when the newborn begins to be progressively colonized by microorganisms until its definitive microbiota is established ^32^. This is exactly what happens, for example, with the gut microbiome of infants, whose alpha diversity is consistently found to be lower than in adults ^33,34^, probably due to the introduction of new diversity from food, which increases with the consumption of foods other than breast milk ^35^. Detailed metagenomic studies, which should include samples collected at multiple time points during childhood and during adulthood, would be needed to determine whether something similar occurs in the nasopharynx. In the case of the present study, we would not be able to detect such differences in diversity between the youngest individuals and the rest of age groups, if they exist, for 2 reasons: (i) because our first age group, which includes the youngest individuals, is very broad and encompasses individuals up to 20 years old, so any differences from infants could be diluted; (ii) because as we were not interested specifically in children, but in broader age groups spanning all stages of life, we excluded children younger than 1 year of age from this study, as their highly changing microbiota in formation could bias the analyses we were interested in. Interestingly, although it is known that there are relevant sex-associated differences in diversity in other anatomical areas such as the gut, and that such differences are probably due to the different levels of sex hormones between both sexes ^36^, we did not observe any significant diversity differences between females and males, even in the age groups where sex hormones levels should be quite different. It is also curious that, contrary to what happens in the gut where the alpha diversity of the microbiota decreases with aging ^30^, we did not observe a similar reduction in diversity in the nasopharynx of older individuals. This difference between both anatomical areas might be explained by the fact that decreased microbial diversity of the gut in older subjects is associated with chronological age, number of concomitant diseases, number of medications used, increasing coliform numbers, and changes in diet ^37^, and although chronological age affects both anatomical areas in a similar way, it seems probable that other of these factors might affect the gut microbiota in a more intense way compared to the nasopharyngeal microbiota. In summary, we can say that the diversity of the nasopharyngeal community is very stable throughout life and between sexes.

Our analyses of nasopharyngeal taxonomic composition at the phylum level revealed the existence of sex-associated differences within the age groups including individuals between 21 and 70 years of age, but not in the youngest and oldest people. As previously mentioned for the alpha diversity results, multiple sex-associated differences in taxonomic composition have also been described for the human gut microbiome ^36^, and numerous studies have found evidence to support the idea that levels of sex hormones, such as progesterone ^38^, androgen ^39^, and oestrogen ^40^, regulate its composition ^41,42^. The effects of sex hormones on other microbial niches, such as the human vaginal microbiota, have also recently been demonstrated ^43^. Although it has been shown that oestrogen stimulation (hormone/gender effect) in the upper respiratory tract mucosa could reduce virus virulence by improving both nasal clearance and local immune response ^44^, the relationship between sex hormone levels and the nasopharyngeal microbiota has so far not been directly proven in clinical studies. However, the fact that our results reveal differences between females and males at the phylum level in all the age groups, except for those where differences in sex hormones levels do not exist or are not so strong (> 70 years and 1-20 years old), suggests that sex hormones might be modulating the taxonomic composition of the healthy human nasopharynx.

We might be tempted to think that the microbiota of nearby anatomical sites that are closely related in terms of structure and function should be practically identical. However, the reality seems to be much more complex. A good example of this is that although nasopharynx and nose are adjacent, and previous metagenomic studies comparing the microbiome of both anatomical areas have revealed a clear continuity, there are important differences between the two sites and even niche-specific bacteria ^45^. Thus, while the adult nasal microbiome is overrepresented with *Moraxella, Haemophilus, Neisseria, Streptococcus, Dolosigranulum, Gemella, Granulicatella, Corynebacterium, Propionibacterium, Turicella*, and *Simonsiella* ^46^, our taxonomic composition results at the genus level show that several of those bacterial genera, such as *Granulicatella, Turicella* or *Simonsiella*, do not even account for more than 1% of the nasopharyngeal microbiota in any of the age groups. Furthermore, our analyses of the taxonomic composition at the genus level only found statistically significant relative abundance differences between the different age groups for 11 bacterial genera: *Acinetobacter, Brevundimonas, Dolosigranulum, Finegoldia, Haemophilus, Leptotrichia, Moraxella, Peptoniphilus, Pseudomonas, Rothia*, and *Staphylococcus*. Interestingly, most of the 37 statistically significant differences found between the different age groups for these 11 genera appear when comparing age groups A1 and A6 with the rest of the age groups (18 out of 37 and 16 out of 37, respectively). Therefore, these results reveal that, in terms of relative abundance of bacterial genera, the nasopharyngeal microbiota of the youngest and oldest subjects is more different from that of the other age groups than that of any other age group. Among these age-associated changes, from a clinical perspective it is particularly concerning the fact that *Dolosigranulum*, which is an opportunistic pathogen that causes pneumonia in elderly patients ^47^, is overrepresented in the nasopharynx of individuals over 70 years of age compared to middle-aged subjects. This suggests that the relative abundance of *Dolosigranulum* may be higher in elderly people due to the process of immunosenescence that occurs in them ^48,49^, or that its higher abundance may be due to other unidentified age-related factors. In either case, in both hypothetical cases, the process of immunosenescence would make elderly individuals more susceptible to an opportunistic lung infection caused by *Dolosigranulum*. In the opposite direction, it is paradoxical that *Haemophilus*, that causes pneumonia mainly in elderly people ^50^, is underrepresented precisely in individuals over 70 years of age. This suggests that people with high relative abundance of *Haemophilus* die prematurely, or that their lower relative abundance in elderly subjects is due to other unidentified age-related factors and that older individuals are much more susceptible to opportunistic infections caused by this bacterium, probably due to the previously mentioned process of immunosenescence. Something similar could be said for *Rothia*, as its relative abundance also decreases drastically in people over 70 years of age while it is known to cause pneumonia mostly in aged individuals ^51^. It is worth noting that, regardless of their relative abundance or whether they show statistically significant differences between age or sex groups, those bacterial genera that are present in most individuals or whose frequency of appearance changes drastically throughout life could be relevant from a biomedical and ecological point of view. Based on this idea, we highlight *Anaerococcus, Burkholderia, Campylobacter, Delftia, Prevotella, Neisseria, Propionibacterium, Streptococcus, Ralstonia, Sphingomonas* and *Corynebacterium* as candidate bacterial genera that could be playing an important role as they are present in the nasopharynx of most healthy individuals. In addition, we propose *Faecalibacterium, Stenotrophomonas* and *Phascolarctobacterium* as candidate bacterial genera that could be playing a relevant role, as their frequency of appearance in the nasopharynx of healthy subjects increases progressively throughout life, and *Aggregatibacter, Gemella* and *Fusobacterium* because their frequency of appearance in the nasopharynx decreases drastically and progressively as healthy people age. Elucidating the biomedical relevance of all these bacterial genera which are part of the healthy nasopharyngeal microbiota and determining their potential involvement in health and disease at different stages of life, is certainly an exciting topic for future work.

Although multiple studies have analyzed the microorganisms present in the nasopharynx in different contexts before this work, the composition of the healthy and mature human nasopharyngeal microbiota was largely unknown since i) most studies focused on children or elderly people, ii) confounding factors such as external drivers that alter it are not well known to date, and iii) a focus is generally done on its variation in diseases. With this work, we fill this important gap in knowledge. However, further research will be necessary to elucidate how the nasopharyngeal taxonomic composition as well as the age- and sex-associated changes described here, affect the susceptibility of certain individuals to infectious diseases. Studying the case of the elderly people in detail will be particularly interesting from a biomedical and clinical perspective, since their nasopharyngeal microbiota is significantly different from that of younger subjects, and they are known to be much more susceptible to multiple infectious diseases, most notably COVID-19 ^52^. Therefore, it would not be too bold to hypothesize that there may be some correlation between the taxonomic composition in the nasopharynx of the elderly and their increased susceptibility to COVID-19, but this will be a challenge for future metagenomic studies that should include different age groups, both sexes, and patients infected with SARS-CoV-2 who have developed the disease with different severity.

## METHODS

### Sample selection, collection, and classification

We randomly selected 120 nasopharyngeal samples from a cohort of 6,354 healthy subjects belonging to the Health Area I of the Region of Murcia (Spain) and who voluntarily gave their samples for diagnostic purposes. Nasopharyngeal swabs were obtained by approaching the nasopharynx transnasally between August 27, 2020, and September 8, 2020, and the Microbiology Service of the Hospital Clínico Universitario Virgen de la Arrixaca (Murcia, Spain) used polymerase chain reaction (PCR) to verify that all the samples were negative for SARS-CoV-2 infection. The kit used for the PCR test was Novel Coronavirus (2019-nCoV) Real Time Multiplex RT-PCR kit (Detection for 3 Genes), manufactured by Shanghai ZJ Bio-Tech Co., Ltd. (Liferiver) and the CFX96 Touch Real-Time PCR Detection System (BioRad).

To facilitate the study of age-associated changes in the nasopharyngeal microbiota throughout life, the samples were binned into six age groups (Table S1). In addition, within each age group, half of the samples were from males and the other half from females in order to study the possible sex-associated changes in the nasopharyngeal microbiota at different life stages (Table S1).

### Amplification, library preparation, and sequencing

Bacterial identification was performed by sequencing the 16S rRNA gene’s hypervariable regions. The 16S rRNA gene was amplified by multiplex PCR using Ion Torrent 16S Metagenomics kit (Ion Torrent, Thermo Fisher Scientific Inc.), using two sets of primers, which targets regions V2, V4, and V8, and V3, V6-7, and V9, respectively. Amplification was carried out in a SimpliAmp thermal cycler (Applied Biosystems) running the following program: denaturation at 95°C for 10 min, followed by a 3-step cyclic stage consisting of 25 cycles of denaturation at 95°C for 30 s, annealing at 58°C for 30 s, and extension at 72°C for 20 s; at the end of this stage, the program concludes with an additional extension period at 72°C for 7 min and the reaction is stopped by cooling at 4°C. The resulting amplicons were tested by electrophoresis through 2% agarose gels in tris-acetate-EDTA (TAE) buffer, purified with AMPure® XP Beads (Beckman Coulter, Inc, Atlanta, GA, USA), and quantified using QubitTM dsDNA HS Assay Kit in a Qubit 3 fluorometer (Invitrogen, Thermo Fisher).

A library was generated from each sample using the Ion Plus Fragment Library Kit (Ion Torrent), whereby each library is indexed by ligating Ion Xpress ™ Barcode Adapters (Ion Torrent) to the amplicons. Libraries were purified with AMPure® XP Beads and quantified using the Ion Universal Library Quantitation Kit (Ion Torrent) in a QuantStudio 5 Real-Time PCR Instrument (Applied Biosystems). The libraries were then pooled and clonally amplified onto Ion Sphere Particles (ISPs) by emulsion PCR in an Ion OneTouch™ 2 System (Ion Torrent) according to the manufacturer’s instructions. Sequencing of the amplicon libraries was carried on an Ion 530™ Kit (Ion Torrent) on an Ion S5™ System (Ion Torrent). After sequencing, the individual sequence reads were filtered by the Torrent Suite ™ Software v5.12.1 to remove low quality and polyclonal sequences.

### Bioinformatics and statistical analysis

The obtained sequences were analyzed and annotated with the Ion Reporter 5.18.2.0 software using the 16S rRNA Profiling workflow 5.18. Clustering into OTUs and taxonomic assignment were performed based on the Basic Local Alignment Search Tool (BLAST) using two reference libraries, MicroSEQ® 16S Reference Library v2013.1 and the Greengenes v13.5 database. For an OTU to be accepted as valid, at least ten reads with an alignment coverage ≥ 90% between hit and query were required. Identifications were accepted at the genus and species level with sequence identity > 97% and > 99%, respectively. Annotated OTUs were then exported for analysis with R (v.4.1.2) (https://www.R-project.org/), where data were converted to phyloseq object ^53^ and abundance bar plots were generated. Data were converted to DESeq2 object ^54^, that uses a generalized linear model based on a negative binomial distribution, to calculate differential abundance between groups. Principal component analysis (PCA) was performed using the abundance data of the different taxa for each individual. Alpha diversity was estimated based on Chao1, Shannon and Inverse-Simpson indices by using the phyloseq package. To test for statistically significant differences between pairwise groups in alpha diversity, the non-parametric Wilcoxon test was used. Frequency of appearance was obtained by calculating the percentage of individuals in each age group in which that taxon occurs. The bar plots aggregated by groups (age and/or gender) show the aggregated relative abundance (sum of relative abundances). Krona charts, that aid in the estimation of relative abundances even within complex metagenomic classifications, were generated as previously described ^55^. All the other graphs were generated with the R package ggplot2 version 3.3.3. ^56^.

## Data Availability

Raw sequencing data of all 16S rRNA sequences, metadata, and abundance tables are available at the open access repository Figshare under the accession numbers 10.6084/m9.figshare.19785991, 10.6084/m9.figshare.19786147, and 10.6084/m9.figshare.19785991, respectively.

## Acknowledgements

We thank I. Fuentes, P. Martinez for their excellent technical assistance, Dr. Anabel Antón for 16S rRNA sequencing, and the staff of the Microbiology Service of HCUVA for sample collection and processing.

## Authors’ contributions

The authors offer the following declarations about their contributions: Conceived and designed the experiments: SC, MLC, VM. Performed the experiments: SC, FP-S. Analyzed the data: SC, FP-S, SDT, AE, MLC, VM. Provided essential samples: AM-C. Wrote the paper: SC, VM.

## Funding

This work was supported by the grant 00006/COVI/20 to VM and MLC funded by Fundación Séneca-Murcia, the Saavedra Fajardo contract to SC funded by Fundación Séneca-Murcia, the Juan de la Cierva-Incorporación contract to SDT funded by Ministerio de Ciencia y Tecnología/AEI/FEDER. The funders had no role in the study design, data collection and analysis, decision to publish, or preparation of the manuscript.

## Ethics approval and consent to participate

All procedures in this work were carried out following the principles expressed in the Declaration of Helsinki, as well as in all the other applicable international, national, and/or institutional guidelines for the use of samples and data, and have been approved by the Comité de Ética de la Investigación (CEIm) at Hospital Clínico Universitario Virgen de la Arrixaca (protocol number 2020-10-12-HCUVA – Effects of aging in the susceptibility to SARS-CoV-2). Nasopharyngeal swabs were collected for diagnosis of SARS-CoV-2 infection before this study was conceived, without the need of any informed consent as the collection procedure was non-invasive and risk-free. However, when the COVID-19 pandemic spread out of control, samples started to be kept at the Microbiology Service instead of destroyed after diagnosis as it was considered that they might be extremely relevant for research. This, together with the facts that i) the retrospective use of these samples did not affect donor health or treatment at all, ii) all data has been treated anonymously, and iii) movement was limited due to the exceptional circumstances of the pandemic meant that it was not possible to obtain informed consents for the use of these samples in research. Moreover, none of the subjects expressly objected to their samples being used for research.

## Competing interests

The authors declare no competing interests.

## Supplementary figure legends

**Figure S1.**
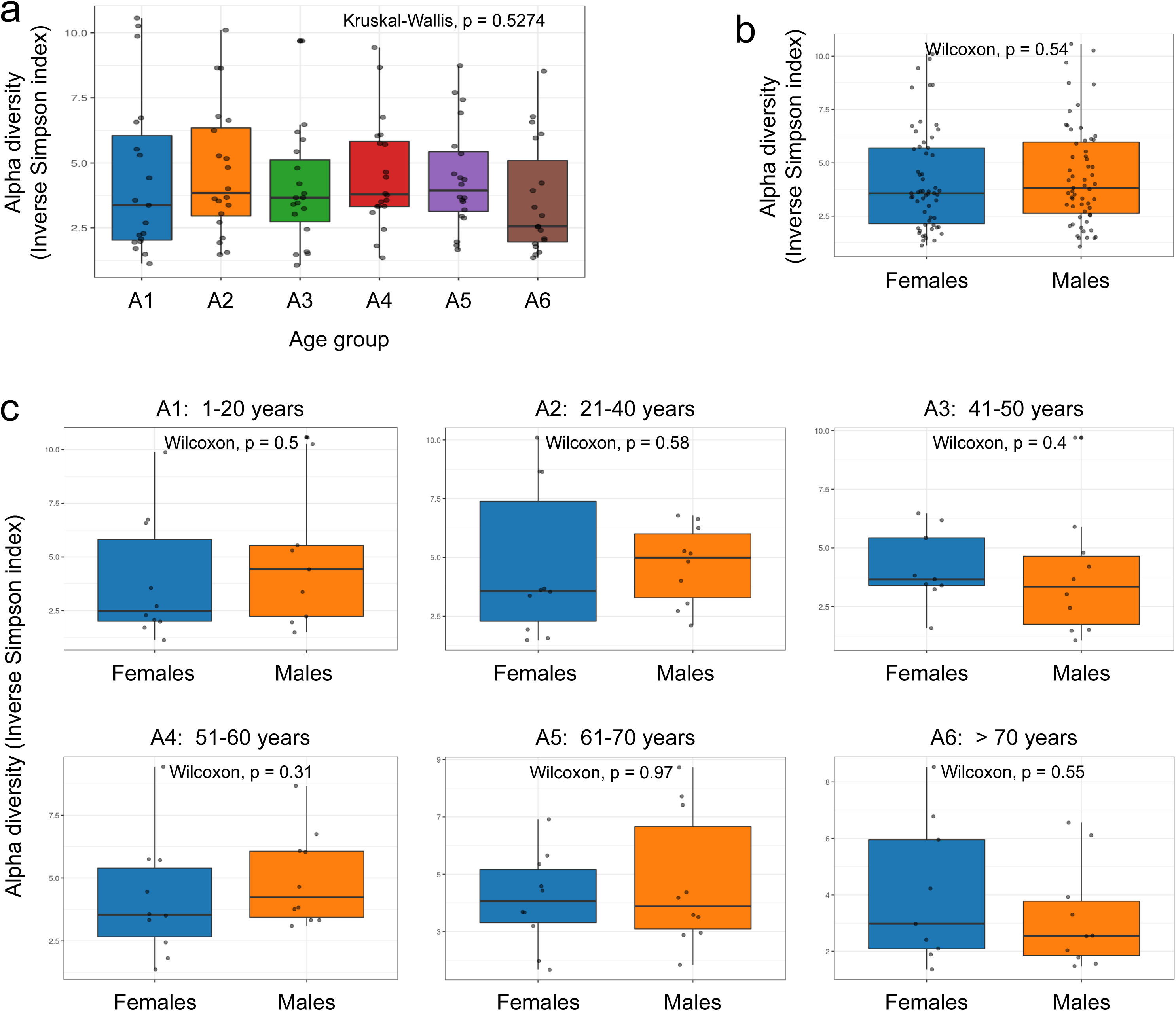
Comparison of alpha diversity parameters across the age and sex groups studied. Box-whisker plots of the alpha diversity inverse Simpson index and its comparison using the Kruskal-Wallis test among the different age groups established for this study (**a**), and the Wilcoxon signed-rank test between females and males (**b-c**). Each sample is represented by one dot.

**Figure S2.**
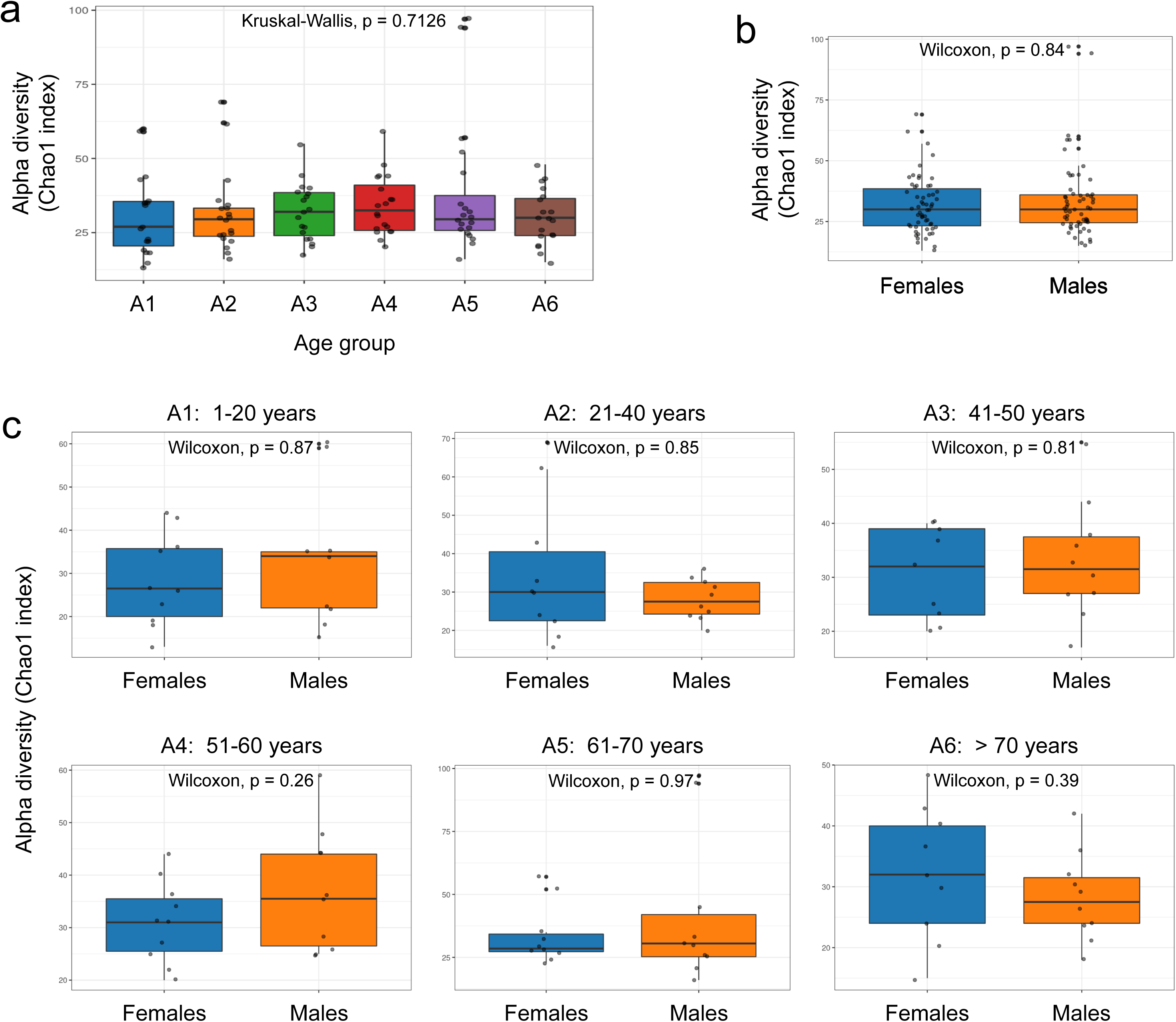
Comparison of alpha diversity parameters across the age and sex groups studied. Box-whisker plots of the alpha diversity Chao1 index and its comparison using the Kruskal-Wallis test among the different age groups established for this study (**a**), and the Wilcoxon signed-rank test between females and males (**b-c**). Each sample is represented by one dot.

**Figure S3.**
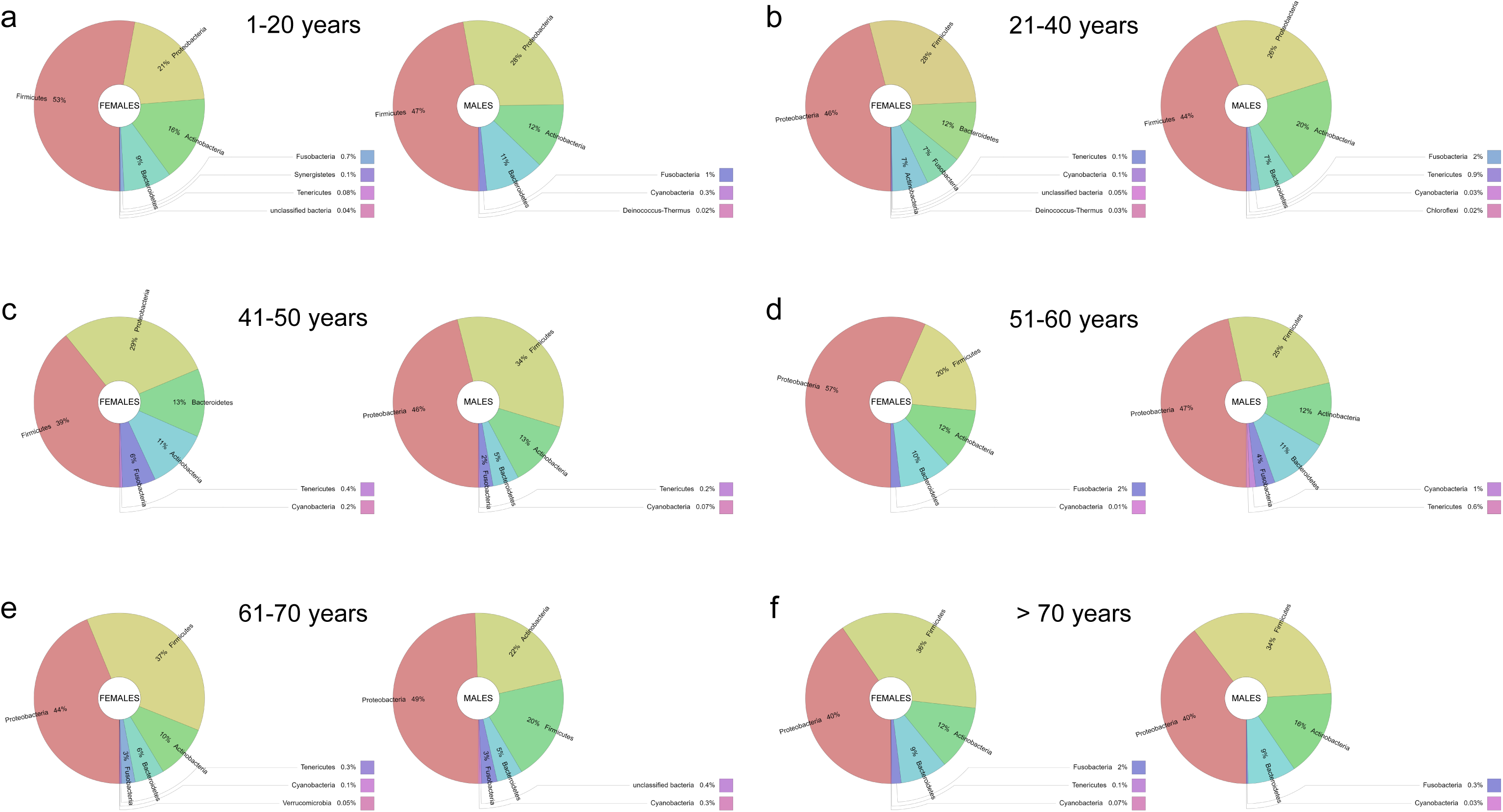
Taxonomic composition and age- and sex-associated metagenomic changes in the nasopharynx of healthy people. **a-f** Krona charts showing bacterial community composition at phylum level in the indicated age and sex groups.

**Figure S4.**
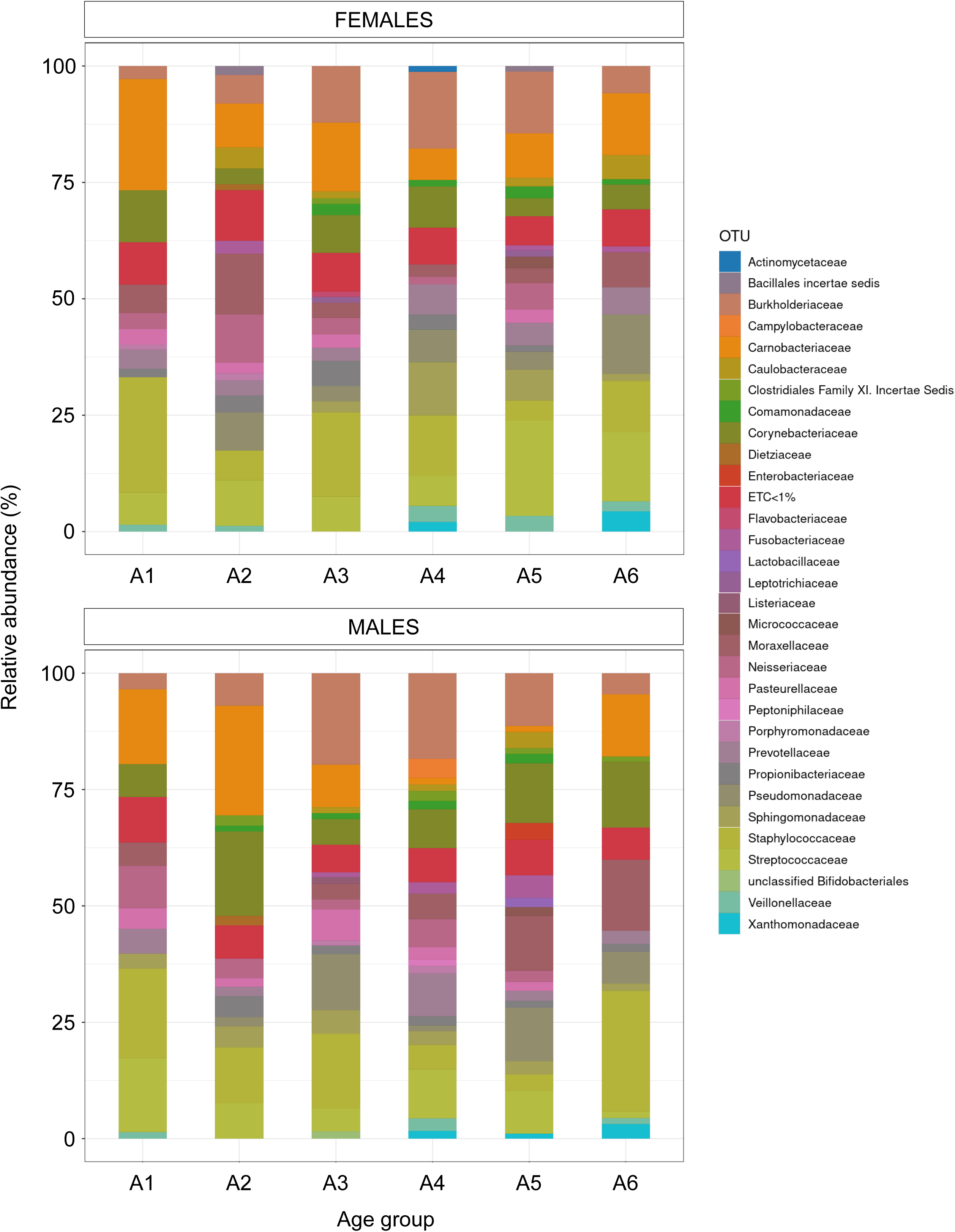
Taxonomic composition and age- and sex-associated metagenomic changes at the family level in the nasopharynx of healthy people. Stacked bar charts showing the relative abundance (%) of bacterial families in the indicated age groups and separately by sex. For clarity, only the bacterial families with average abundance > 1% at each age group are shown.

**Figure S5.**
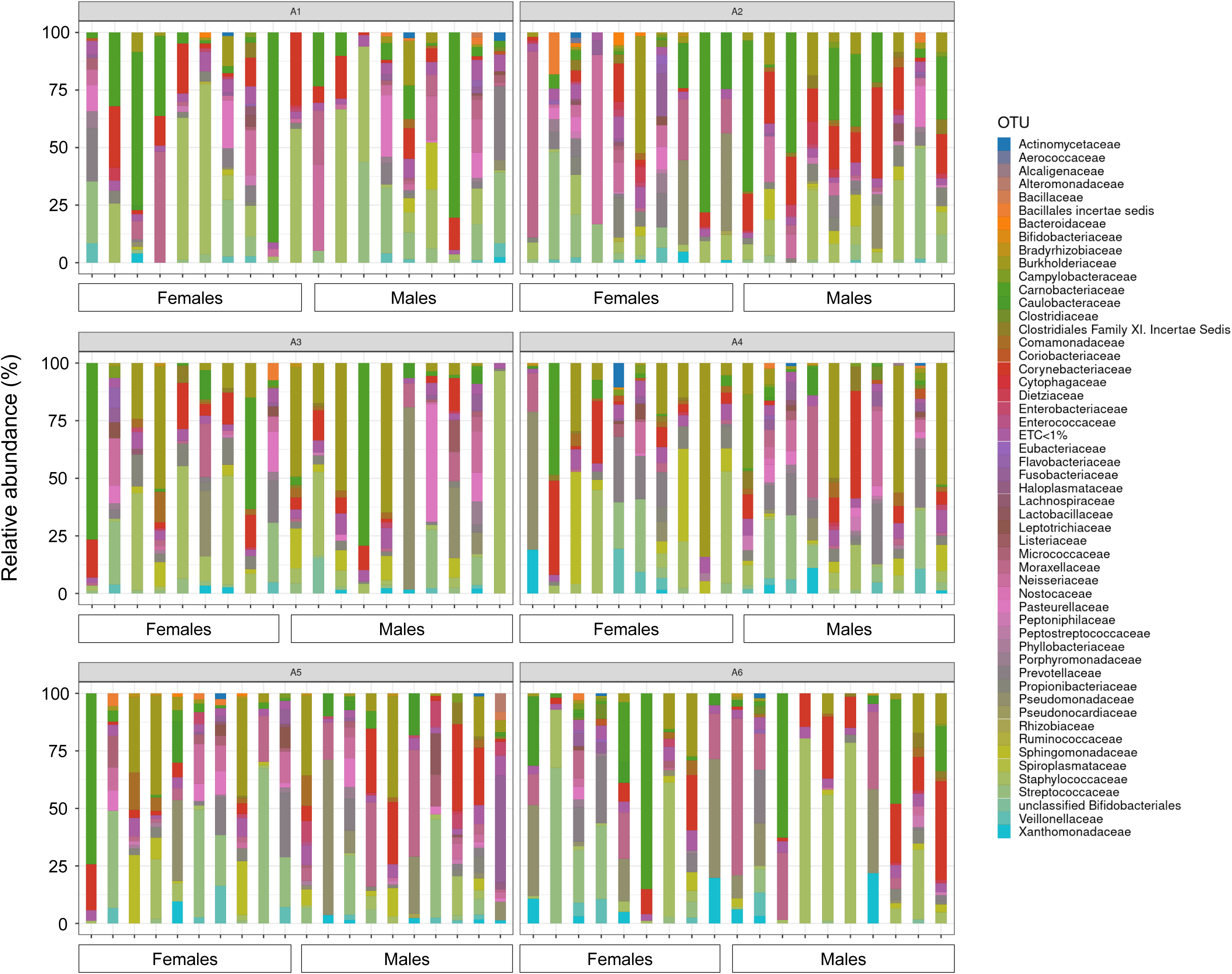
Taxonomic composition and age- and sex-associated metagenomic changes at the family level in the nasopharynx of each individual included in this study. Stacked bar charts showing the relative abundance (%) of bacterial families, in the indicated age groups and separately by sex, of each subject included in this study. For clarity, only the bacterial families with average abundance > 1% at each age group are shown.

**Figure S6.**
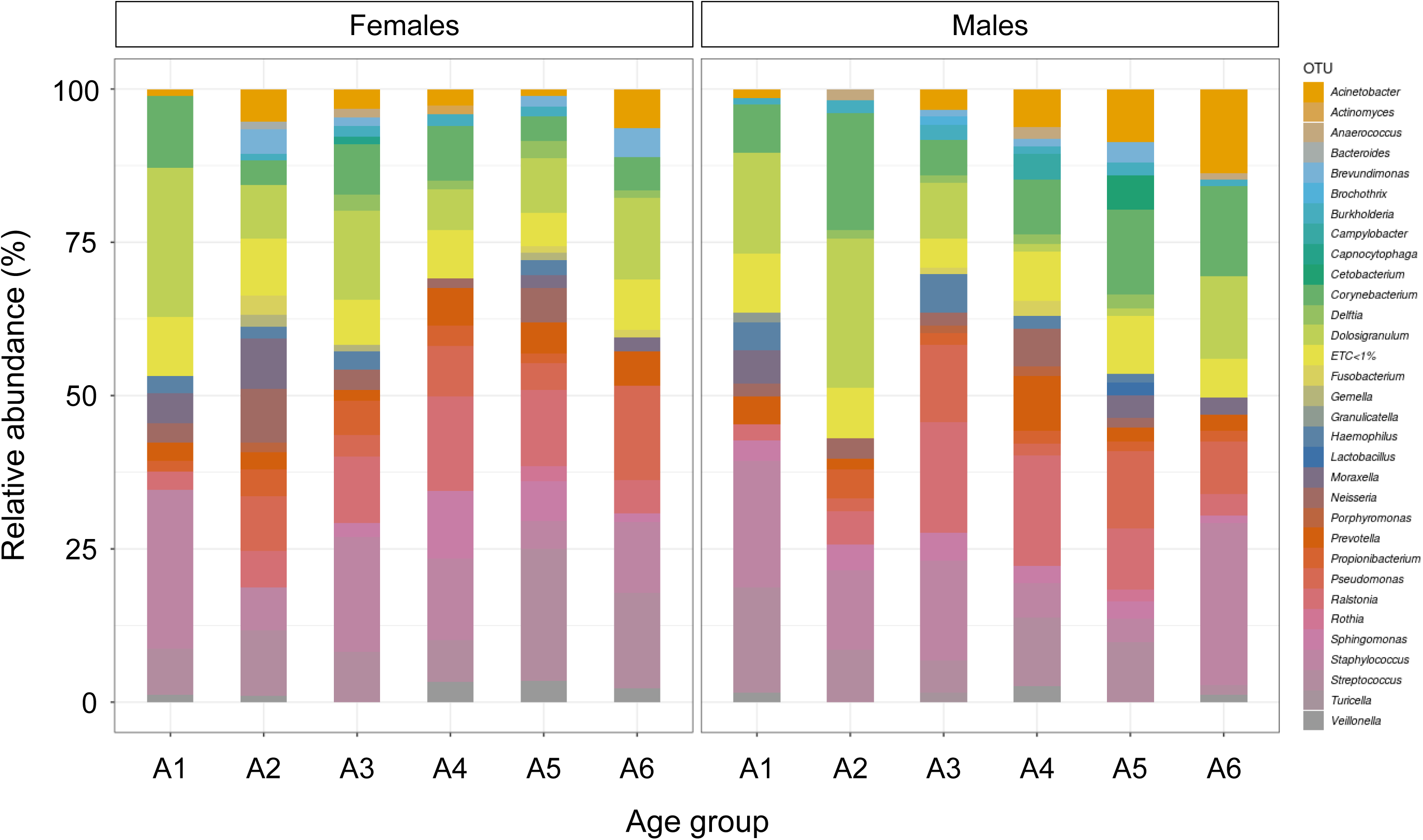
Taxonomic composition and age- and sex-associated metagenomic changes at the genus level in the nasopharynx of healthy people. Stacked bar charts showing the relative abundance (%) of bacterial genera in the indicated age groups and separately by sex. For clarity, only the bacterial genera with average abundance > 1% at each age group are shown.

**Figure S7.**
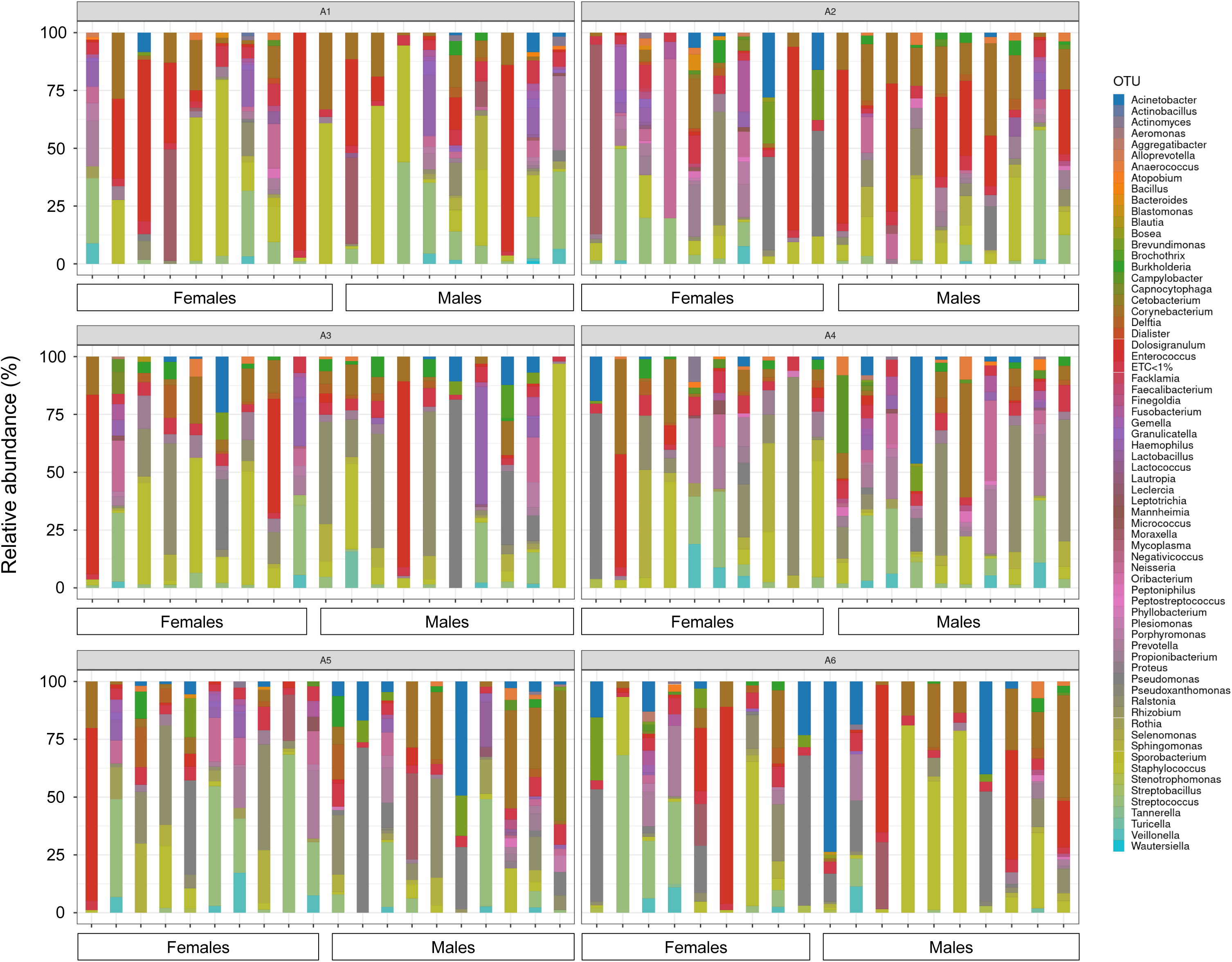
Taxonomic composition and age- and sex-associated metagenomic changes at the genus level in the nasopharynx of each individual included in this study. Stacked bar charts showing the relative abundance (%) of bacterial genera, in the indicated age groups and separately by sex, of each subject included in this study. For clarity, only the bacterial genera with average abundance > 1% at each age group are shown.

**Figure S8.**
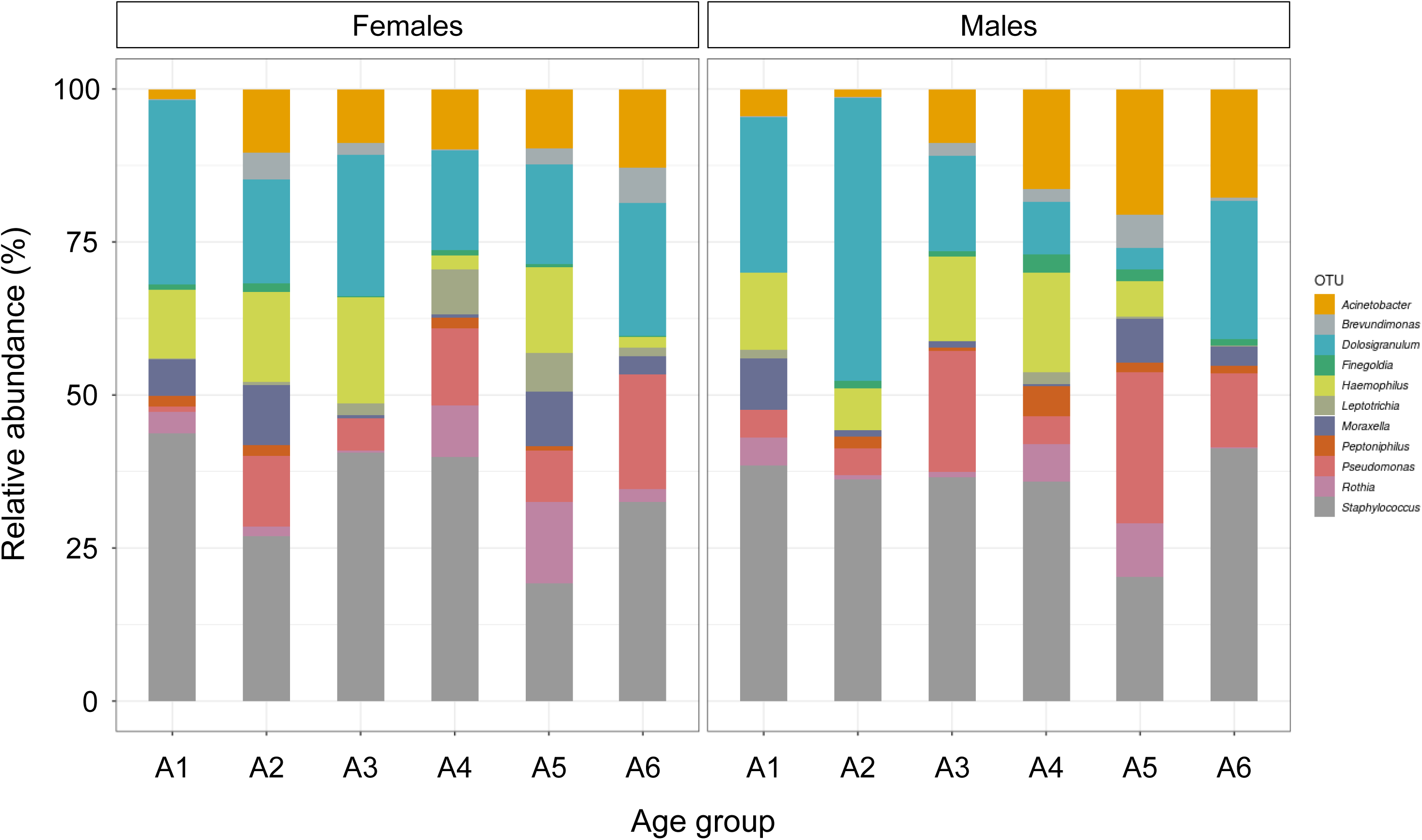
Taxonomic composition and age- and sex-associated metagenomic changes at the genus level in the nasopharynx of healthy people. Stacked bar charts showing the relative abundance (%) of the 11 selected bacterial genera indicated, in the indicated age groups and separately by sex.

**Figure S9.**
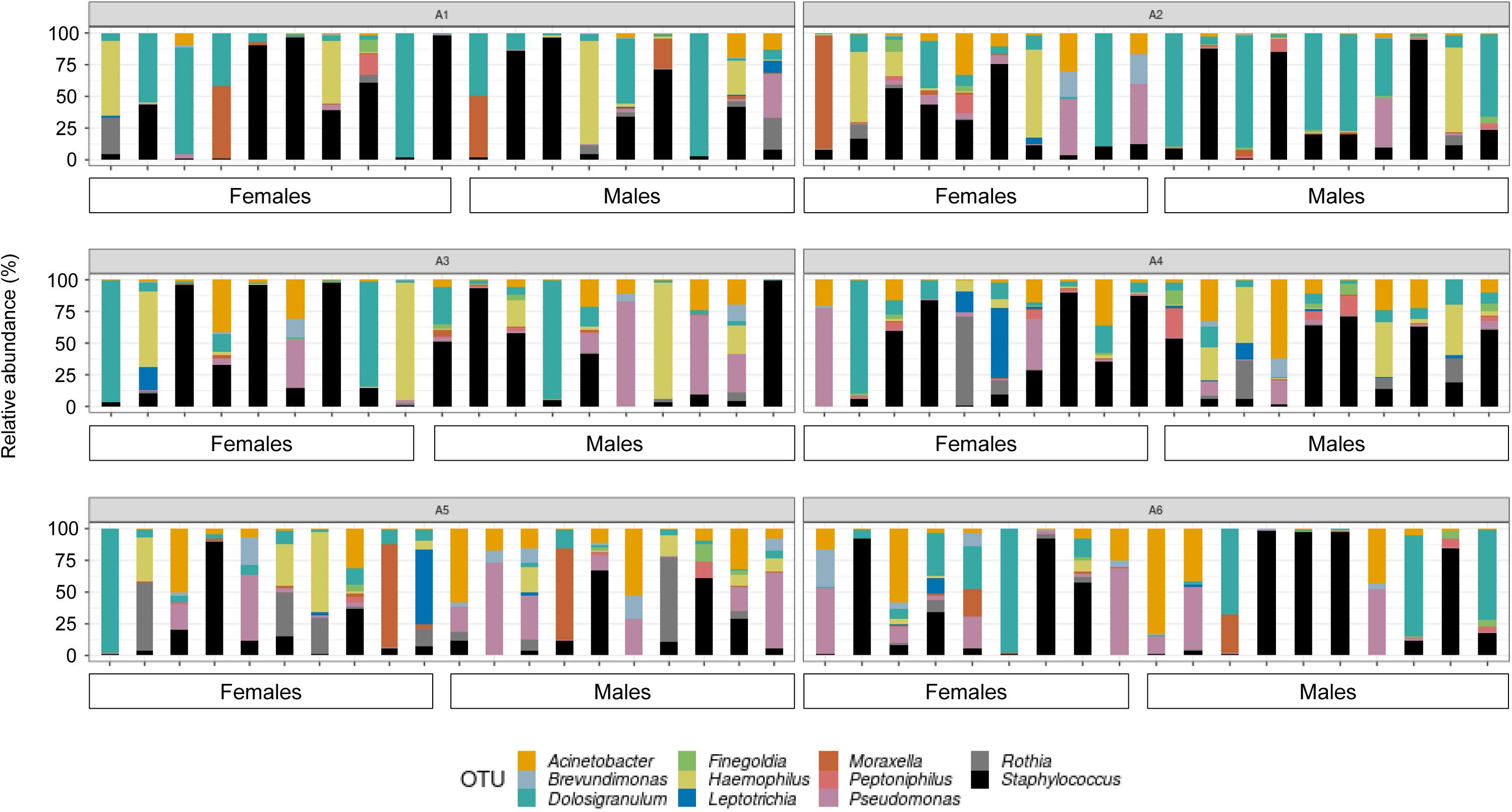
Taxonomic composition and age- and sex-associated metagenomic changes at the genus level in the nasopharynx of each individual included in this study. Stacked bar charts showing the relative abundance (%) of the 11 selected bacterial genera indicated, in the indicated age groups and separately by sex, of each subject included in this study. Note that the rest of bacterial genera are not taken into consideration in this analysis.

**Figure S10.**
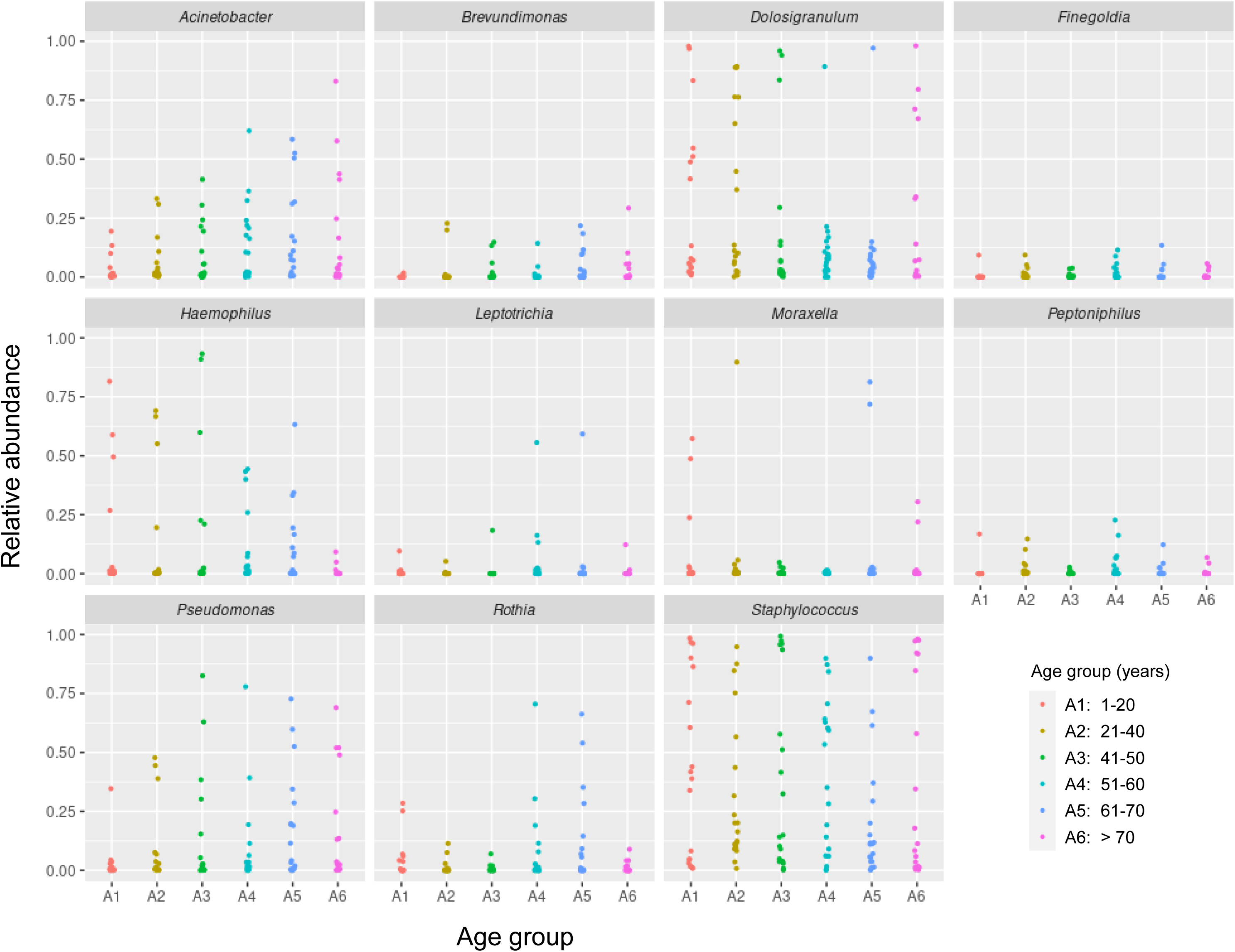
Relative abundance of the 11 bacterial genera which present significant differences between age groups in each individual. Relative abundance of the selected 11 bacterial genera indicated in each subject included in this study, in the indicated age groups. Each sample is represented by one dot, colored according to age.

**Table S1.** Nasopharyngeal exudate samples collected from healthy (not infected with SARS-CoV-2) males and females in each age group

| Age group | Sex | Samples |
| --- | --- | --- |
| A1: 1-20 years | *Female* | 10 |
|  | *Male* | 9 |
| A2: 21-40 years | *Female* | 10 |
|  | *Male* | 10 |
| A3: 41-50 years | *Female* | 10 |
|  | *Male* | 10 |
| A4: 51-60 years | *Female* | 10 |
|  | *Male* | 10 |
| A5: 61-70 years | *Female* | 10 |
|  | *Male* | 10 |
| A6: >70 years | *Female* | 10 |
|  | *Male* | 10 |
| Total | *Female* | 60 |
|  | *Male* | 59 |

**Table S2.** Summary of the statistical analysis of the relative abundance differences among the 6 age groups established in this study. Only the statistically significant differences (adjusted p-value < 0.05) for the 11 bacterial genera which present such differences are shown

| Genus | baseMean | log2FoldChange | lfcSE | stat | Adjusted p-value | Age groups compared |
| --- | --- | --- | --- | --- | --- | --- |
| *Acinetobacter* | 848,877 | -2,463 | 0,664 | -3,71 | 0,009 | A1_A6 |
|  |  | -2,866 | 0,655 | -4,374 | 0,003 | A2_A6 |
|  |  | 2,378 | 0,655 | 3,629 | 0,02 | A3_A4 |
|  |  | -3,688 | 0,655 | -5,628 | 0 | A4_A6 |
|  |  | -2,338 | 0,655 | -3,57 | 0,019 | A5_A6 |
| *Brevundimonas* | 129,952 | -5,191 | 1,366 | -3,799 | 0,01 | A1_A2 |
|  |  | -5,772 | 1,383 | -4,173 | 0,002 | A1_A6 |
| *Dolosigranulum* | 16684,133 | 5,033 | 0,977 | 5,154 | 0 | A1_A4 |
|  |  | 3,581 | 0,964 | 3,714 | 0,021 | A2_A4 |
|  |  | 4,244 | 0,977 | 4,346 | 0,001 | A3_A4 |
|  |  | -4,553 | 0,977 | -4,662 | 0 | A4_A6 |
| *Finegoldia* | 50,796 | -5,379 | 1,527 | -3,523 | 0,022 | A1_A2 |
| *Haemophilus* | 450,095 | 3,981 | 0,961 | 4,141 | 0,002 | A1_A6 |
|  |  | 4,733 | 0,961 | 4,925 | 0 | A3_A6 |
|  |  | 4,461 | 0,949 | 4,701 | 0 | A5_A6 |
| *Leptotrichia* | 21,087 | 8,756 | 2,111 | 4,148 | 0,005 | A1_A3 |
|  |  | -10,723 | 2,08 | -5,156 | 0 | A3_A4 |
|  |  | -10,303 | 2,08 | -4,953 | 0 | A3_A5 |
|  |  | -7,858 | 2,112 | -3,72 | 0,021 | A3_A6 |
| *Moraxella* | 90,658 | 3,938 | 0,991 | 3,973 | 0,004 | A1_A4 |
| *Peptoniphilus* | 55,534 | -7,681 | 1,739 | -4,416 | 0,001 | A1_A2 |
|  |  | -7,135 | 1,739 | -4,103 | 0,003 | A1_A4 |
|  |  | -6,649 | 1,761 | -3,776 | 0,008 | A1_A6 |
| *Pseudomonas* | 2779,023 | -4,48 | 0,933 | -4,799 | 0 | A1_A2 |
|  |  | -3,823 | 0,945 | -4,044 | 0,005 | A1_A3 |
|  |  | -5,261 | 0,945 | -5,566 | 0 | A1_A6 |
|  |  | 3,676 | 0,921 | 3,994 | 0,014 | A2_A4 |
|  |  | -4,458 | 0,933 | -4,781 | 0 | A4_A6 |
| *Rothia* | 115,404 | -5,551 | 1,281 | -4,332 | 0,003 | A2_A5 |
|  |  | -4,724 | 1,294 | -3,65 | 0,027 | A3_A5 |
|  |  | 5,027 | 1,296 | 3,878 | 0,007 | A5_A6 |
| *Staphylococcus* | 8272,167 | 2,477 | 0,721 | 3,434 | 0,025 | A1_A2 |
|  |  | 2,635 | 0,73 | 3,608 | 0,021 | A1_A3 |
|  |  | 3,539 | 0,721 | 4,908 | 0 | A1_A4 |
|  |  | 4,078 | 0,721 | 5,656 | 0 | A1_A5 |
|  |  | -2,865 | 0,721 | -3,974 | 0,004 | A4_A6 |
|  |  | -3,405 | 0,721 | -4,722 | 0 | A5_A6 |

## Notes

### Competing Interest Statement

The authors have declared no competing interest.

### Funding Statement

This work was supported by the grant 00006/COVI/20 to VM and MLC funded by Fundacion Seneca-Murcia, the Saavedra Fajardo contract to SC funded by Fundacion Seneca-Murcia, the Juan de la Cierva-Incorporacion contract to SDT funded by Ministerio de Ciencia y Tecnologia/AEI/FEDER. The funders had no role in the study design, data collection and analysis, decision to publish, or preparation of the manuscript.

### Author Declarations

All procedures in this work were carried out following the principles expressed in the Declaration of Helsinki, as well as in all the other applicable international, national, and/or institutional guidelines for the use of samples and data, and have been approved by the Comite de Etica de la Investigacion (CEIm) at Hospital Clinico Universitario Virgen de la Arrixaca (protocol number 2020-10-12-HCUVA - Effects of aging in the susceptibility to SARS-CoV-2).

